# Perceptual and cognitive effects of focal tDCS of auditory cortex in tinnitus

**DOI:** 10.1101/2024.01.31.24302093

**Authors:** Amber M. Leaver

## Abstract

**OBJECTIVES:** Noninvasive brain stimulation continues to grow as an effective, low-risk way of improving the symptoms of brain conditions. Transcranial direct current stimulation (tDCS) is particularly well-tolerated, with benefits including low cost and potential portability. Nevertheless, continued study of perceptual and cognitive side effects is warranted, given the complexity of functional brain organization. This paper describes the results of a brief battery of tablet-based tasks used in a recent pilot study of auditory-cortex tDCS in people with chronic tinnitus.

**METHODS:** Volunteers with chronic tinnitus (n=20) completed two hearing tasks (pure-tone thresholds, Words In Noise) and two cognitive tasks (Flanker, Dimension Change Card Sort) from the NIH Toolbox. Volunteers were randomized to active or sham 4×1 Ag/AgCl tDCS of auditory cortex, and tasks were completed immediately before and after the first tDCS session, and after the fifth/final tDCS session. Statistics included linear mixed-effects models for change in task performance over time.

**RESULTS:** Before tDCS, performance on both auditory tasks was highly correlated with clinical audiometry, supporting the external validity of these measures (r^2^>0.89 for all). Although overall auditory task performance did not change after active or sham tDCS, detection of right-ear Words in Noise stimuli modestly improved after five active tDCS sessions (t(34)=-2.07, p=0.05). On cognitive tasks, reaction times were quicker after sham tDCS, reflecting expected practice effects (e.g., t(88)=3.22, p=0.002 after 5 sessions on Flanker task). However, reaction times did not improve over repeated sessions in the active group, suggesting that tDCS interfered with learning these practice effects.

**CONCLUSIONS:** Repeated sessions of auditory-cortex tDCS does not appear to adversely affect hearing or cognition, but may modestly improve hearing in noisy environments and interfere with some types of motor learning. Low-burden cognitive/perceptual test batteries could be a powerful way to identify adverse effects and new treatment targets in brain stimulation research.

## INTRODUCTION

Noninvasive brain stimulation (NIBS) continues to emerge as an effective method to improve symptoms in various conditions, including depression, obsessive-compulsive disorder, and addiction.^1–6^ When following recommended guidelines, NIBS methods also typically avoid side effects beyond discomfort during the stimulation session.^7,8^ Transcranial direct current stimulation (tDCS) is particularly well-tolerated,^7^ and uses simple, affordable equipment that can be used at home with clinical supervision.^9^ However, given the brain’s complexity and the relatively diffuse nature of NIBS stimulation, continued study of perceptual and cognitive side effects is warranted to ensure continued safety and to identify potential additional uses for tDCS and other NIBS methods. Brief cognitive/perceptual batteries using tablet-based applications may be a way to achieve this while minimizing volunteer and experimenter burden.

A recent pilot mechanistic trial tested the effects of focal tDCS targeting auditory cortex on brain function and tinnitus symptoms in people with chronic tinnitus.^10^ As an exploratory part of this study, volunteers also completed a brief 20-minute battery taken from the NIH Toolbox^11^ to test for potential auditory-perceptual and cognitive side effects after stimulation. Two auditory tasks were selected similar to tests used in clinical audiometry, including pure-tone and speech-in-noise detection tasks.^12^ Volunteers also completed two cognitive tasks, the Flanker Task testing response inhibition and the Dimension Change Card Sort task testing executive function (task switching).^13^ The exploratory hypothesis was that auditory-cortex tDCS may improve speech-in-noise perception given previous studies linking lateral and posterior superior temporal cortex to speech perception,^14–18^ but would not change pure-tone thresholds reflecting peripheral auditory function. Cognitive tasks were chosen as control tasks, and were not expected to be affected by auditory-cortex tDCS. Relationships between clinical audiometry and NIH Toolbox auditory measures were also assessed to confirm the validity of these NIH Toolbox tasks. This paper reports the results of these exploratory analyses using previously unreported data from the original pilot trial.^10^

## MATERIALS AND METHODS

### Subjects

This study was carried out in accordance with the Declaration of Helsinki and with approval of the Northwestern University Institutional Review Board, and all participants gave informed written consent before enrollment. Additional methodological details and results for the main study have been published previously, which meets clinical trials reporting requirements;^10^ only details directly relevant to this ancillary study are reported here. Participants (n=20) had chronic tinnitus for at least one year that was present (perceived) when consciously attended to for at least 50% of awake time, and intruded at least 10% of awake time, over the 12 months before screening. Participants also reported having discussed tinnitus symptoms with a physician or audiologist to rule out obvious physical or neurological origin for tinnitus (e.g., acoustic neuroma, Meniere’s Disease, vascular pulsatile tinnitus). Standard safety considerations for MRI and tDCS were exclusionary, including the presence of incompatible or unspecified implants, significant head injury, claustrophobia, scalp injury or other skin conditions, concurrent use of decongestants, antihistamines, benzodiazepines or other anticonvulsants, and anti-psychotics. Significant or severe developmental, neurological, or psychiatric conditions or substance abuse were also exclusionary, but mild-to-moderate mood or anxiety disorder or symptoms were not exclusionary.

### Study Design

Summary of study schedule for tDCS and Auditory/Cognitive Testing is displayed in **Figure 1A**. Participants were randomly assigned to receive active or sham tDCS scheduled on five consecutive days, and Auditory/Cognitive Tasks were done before the first tDCS session, approximately 30 minutes after first tDCS session, and immediately after the fifth and final tDCS session.

**Figure 1.**
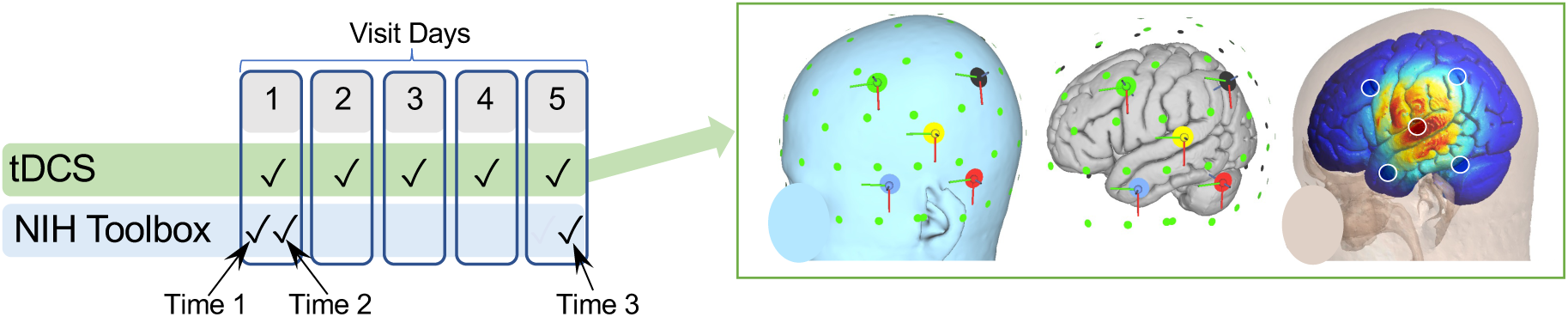
Study methods. Chart at left displays relative timing of tDCS and NIH Toolbox sessions on study visit days. At right, electrode positions are displayed on the reconstructed scalp (left) and cortical surface (middle) on a template head. Small green circles are the 10-10 EEG position grid. At right, electric field magnitude (|E|) is displayed on template head, as estimated using finite element modeling in SimNIBS software.^69^

### TDCS Sessions

Focal tDCS was delivered using a Soterix 1×1 tES device with a 4×1 HD-tDCS conversion box. Electrodes were Ag/AgCl rings placed in custom plastic holders affixed to a spandex cap and filled with Lectron II conductive gel (∼1.5mL/electrode). Current amplitude was 2mA (i.e., +2mA anode, and -0.5mA each cathode), delivered with 30 second linear on- and off-ramps per manufacturer settings. Sessions were 20 minutes in duration; current was constant for active tDCS, and current was ramped up and down at the beginning and end of the 20-minute session for sham stimulation (as programmed by the manufacturer). Paresthesia (e.g., tingling, itching) tends to be greatest during the beginning of the session and during changes in current; transient stimulation in sham programming attempts to equate paresthesia for sham and active conditions in this regard.

Electrode configuration was 4×1,^19^ with center anode positioned over superior temporal (auditory) cortex surrounded by four cathodes (**Figure 1**). Anode position followed the method developed by Langguth et al. ^20^ with small adjustments to accommodate potential differences in head size^21,22^ as described previously.^10^

Each volunteer underwent pure-tone threshold testing at Northwestern Medicine’s Audiology clinic, including standard frequencies and above 8kHz to capture potential high frequency hearing loss. Frequencies tested were: 0.25, 0.5, 1, 2, 3, 4, 6, 8, 10, 12.5, 14, 16, and 20 kHz. Tinnitus assessments included the Tinnitus Functional Index (TFI), Tinnitus Handicap Inventory (THI), and Tinnitus Sample Case History Questionnaire.

Auditory and cognitive tasks were administered via the NIH Toolbox 1.23.4940 using an iPad tablet (8^th^ generation) and Sennheiser Professional HD 280 PRO over-ear headphones for auditory testing. Volunteers were seated at a table in a private room; the room was quiet but not soundproofed. For auditory tasks, volunteers removed hearing aids if present, and volume was manually set to 50% of device output. Note that these volume settings may deviate from the NIH Toolbox instructions for auditory tasks, which instruct administrators to adjust the volume to a midrange setting comfortable to the volunteer. In the current study, the volume was set at the same level for all volunteers to more closely approximate volunteer’s hearing abilities. Apart from this, NIH Toolbox tasks were administered as directed in the Administrator’s Manual for versions 1.23+.

NIH Toolbox Hearing Thresholds Test 7+ v1.0 was administered first, lasting about 9 minutes. In this task, pure-tone stimuli were presented separately to each ear at 500, 1000, 2000, and 4000 Hz. Volunteers were asked to respond via button-press on the tablet when they heard a tone. The threshold for each frequency was the sound level at which the volunteer responded with consistency according to the NIH Toolbox App. Frequencies tested were more limited than clinical diagnostic audiometry, but overlap the range important for speech perception.

NIH Toolbox Words-In-Noise Test Age 6+ v2.1 was administered second and lasted about 6 minutes. In this task, a single-syllable word spoken by a single female speaker was presented simultaneously with multi-speaker babble (noise). The volunteer was asked to repeat the word spoken by the female speaker, and correct responses were recorded by the experimenter using the tablet. The level (amplitude) of the word is varied over seven signal-to-noise ratios (SNR) with respect to the multi-speaker babble. Trials started with the highest SNR, where the target word was 24 SNR higher than the noise, followed by 20, 16, 12, 8, 4, to 0 SNR. There were five trials repeated at each SNR level, and each ear was tested separately. There were two lists of stimuli, one for each ear within a session; the order in which the lists are presented was selected at random by the NIH Toolbox App. So, words are not repeated within each session, but were repeated over repeat sessions in this study. Threshold was calculated in dB SNR by NIH Toolbox for each ear.

NIH Toolbox Flanker Inhibitory Control and Attention Test was administered third. In this task, participants viewed an arrow, flanked on the left and right by arrows that either pointed in the same (congruent) or different (incongruent) direction as the middle arrow. One each trial, volunteers were asked to indicate which direction the middle arrow was pointing via button-press on the tablet using one finger. Between trials, volunteers placed this finger at “home base”, a dot located on the table approximately 3 inches from the base of the tablet. This task had 20 trials total and took about 3 minutes. Computed Score and reaction times (RTs) are reported. Because accuracy is typically high in cognitively healthy groups, RTs are reported for correct trials only. Trials where RT was less than 100 msec or greater than 3 standard deviations above mean RT were omitted from analysis. Thus, 16 trials (9 active group, 7 sham group) were omitted for 11 total volunteers (5 active, 6 sham) over the three sessions; the highest number of trials omitted for any volunteer was 3.

In the NIH Toolbox Dimension Change Card Sort (DCCS) Test, participants were asked to match a visual object located at the bottom of the screen with one of two objects at the top of the screen. In the Shape condition, volunteers matched by object shape (irrespective of color), and in the Color condition, volunteers matched by object color (irrespective of shape). The volunteer was prompted of the current task rule with the word “Shape” or “Color” at the beginning of each trial, and the volunteer responded by touching the matching shape on the tablet screen (returning their finger to “home base” between trials). For this task, the key experimental manipulation was during condition switches from Shape to Color matching rules and vice versa. The degree of slowing (and/or number of errors) on rule Switch trials compared with Repeat rule trials are a measure of executive function. There were 30 trials in this task, which took about 4 minutes. Again, both Computed Score and RTs are reported. Again, trials where RT was less than 100 msec or greater than 3 standard deviations above mean RT were omitted from analysis, along with RTs from incorrect trials. Thus, 39 trials (19 active, 20 sham) were omitted for 20 total volunteers (10 active, 10 sham) over three sessions; the highest number of trials omitted for any volunteer was 4.

### Statistical Analyses

All statistical analyses were completed in R (https://www.r-project.org), including libraries lme4^23^ and lmerTest.^24^ Age, sex, and mean pure-tone audiometric threshold did not vary between groups (**Table 1**); therefore, these were not used as nuisance factors in statistical models. Statistical tests of relevant demographic and clinical variables were done using t or Chi-square tests to test for differences between active and sham groups. Relationships between audiometry results and NIH Toolbox auditory task performance were assessed by calculating Pearson’s correlation coefficients for scores of interest. Because their distributions are well-known to be not normal,^25^ RTs were log-transformed prior to statistical analysis for both cognitive tasks.

**Table 1.**
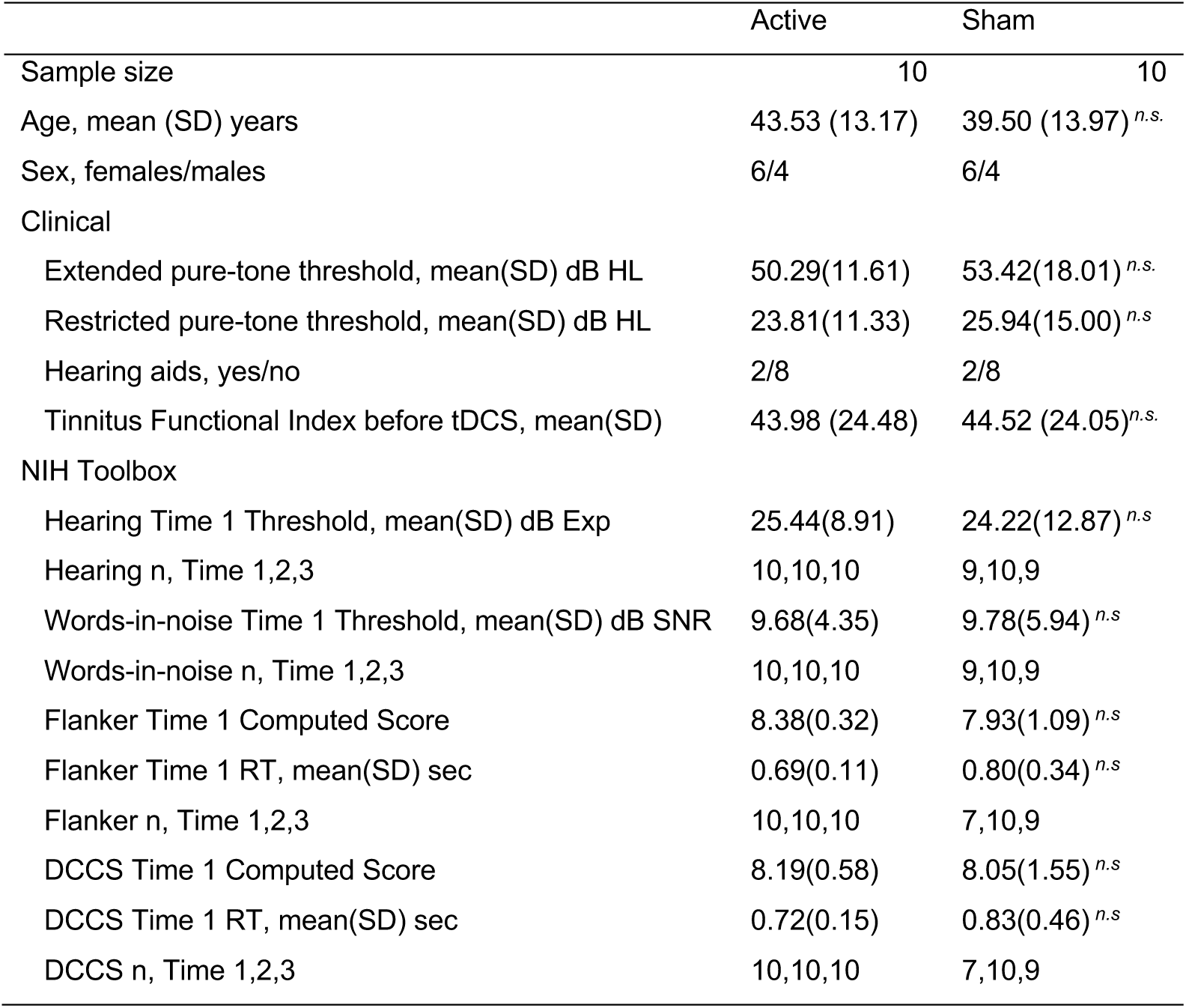
Participant Information.

|  | Active | Sham |
| --- | --- | --- |
| Sample size | 10 | 10 |
| Age, mean (SD) years | 43.53 (13.17) | 39.50 (13.97) *n.s.* |
| Sex, females/males | 6/4 | 6/4 |
| Clinical |  |  |
| Extended pure-tone threshold, mean(SD) dB HL | 50.29(11.61) | 53.42(18.01) *n.s.* |
| Restricted pure-tone threshold, mean(SD) dB HL | 23.81(11.33) | 25.94(15.00) *n.s.* |
| Hearing aids, yes/no | 2/8 | 2/8 |
| Tinnitus Functional Index before tDCS, mean(SD) | 43.98 (24.48) | 44.52 (24.05) *n.s.* |
| NIH Toolbox |  |  |
| Hearing Time 1 Threshold, mean(SD) dB Exp | 25.44(8.91) | 24.22(12.87) *n.s.* |
| Hearing n, Time 1,2,3 | 10,10,10 | 9,10,9 |
| Words-in-noise Time 1 Threshold, mean(SD) dB SNR | 9.68(4.35) | 9.78(5.94) *n.s.* |
| Words-in-noise n, Time 1,2,3 | 10,10,10 | 9,10,9 |
| Flanker Time 1 Computed Score | 8.38(0.32) | 7.93(1.09) *n.s.* |
| Flanker Time 1 RT, mean(SD) sec | 0.69(0.11) | 0.80(0.34) *n.s.* |
| Flanker n, Time 1,2,3 | 10,10,10 | 7,10,9 |
| DCCS Time 1 Computed Score | 8.19(0.58) | 8.05(1.55) *n.s.* |
| DCCS Time 1 RT, mean(SD) sec | 0.72(0.15) | 0.83(0.46) *n.s.* |
| DCCS n, Time 1,2,3 | 10,10,10 | 7,10,9 |

Changes in task performance in active and sham groups were tested using linear mixed models.^23^ The main analyses targeted a time-by-group interaction to identify differences between active and sham groups in change in task performance over time, as well as main effects of time to identify similar changes in task performance over time in both groups (e.g., overall improvements over sessions due to practice effects). In these models, time was a numerical factor, group was a categorical factor (active, sham), and subject was a random factor (random intercepts). Task condition was an additional categorical factor for models analyzing auditory task performance (ear) and cognitive task RTs (congruent/incongruent for Flanker, repeat/switch for DCCS). Presence of interactions and main effects of time were also reported separately for each task condition (Ear, Congruent/Incongruent, Repeat/Switch). In instances where time-by-assignment interactions or main effects of time were p<0.05, pairwise comparisons of baseline and follow-up timepoints/sessions were also reported.

## RESULTS

### Demographics and Tinnitus Outcomes

As reported previously,^10^ volunteers assigned to active or sham tDCS groups did not differ with respect to age, sex, mean pure-tone audiometric threshold, hearing aid use, mood disorder symptoms, age of tinnitus onset, or baseline tinnitus symptom severity (i.e., TFI score, Tinnitus Loudness rating, and Tinnitus Awareness rating; **Table 1**). Groups also did not differ on baseline performance on auditory and cognitive tasks (**Table 1**). Changes in tinnitus severity after tDCS were reported previously, and include modest reductions in Tinnitus Loudness ratings two weeks after active tDCS and in Tinnitus Awareness ratings four weeks after active tDCS.^10^

### Auditory Tasks

Before tDCS, pure-tone thresholds tested in the audiology clinic and using NIH Toolbox were highly correlated, and both pure-tone threshold measures were also highly correlated with Words in Noise (WIN) thresholds measured with NIH Toolbox (**Figure 2**). Correspondence was high for each of the four pure-tone frequencies tested in both clinical and experimental settings (**Figure 2B**) and for averaged threshold (**Figure 2C**). WIN thresholds were also highly correlated with clinical and experimental pure-tone thresholds measured before tDCS (**Figure 2D-E)**.

**Figure 2.**
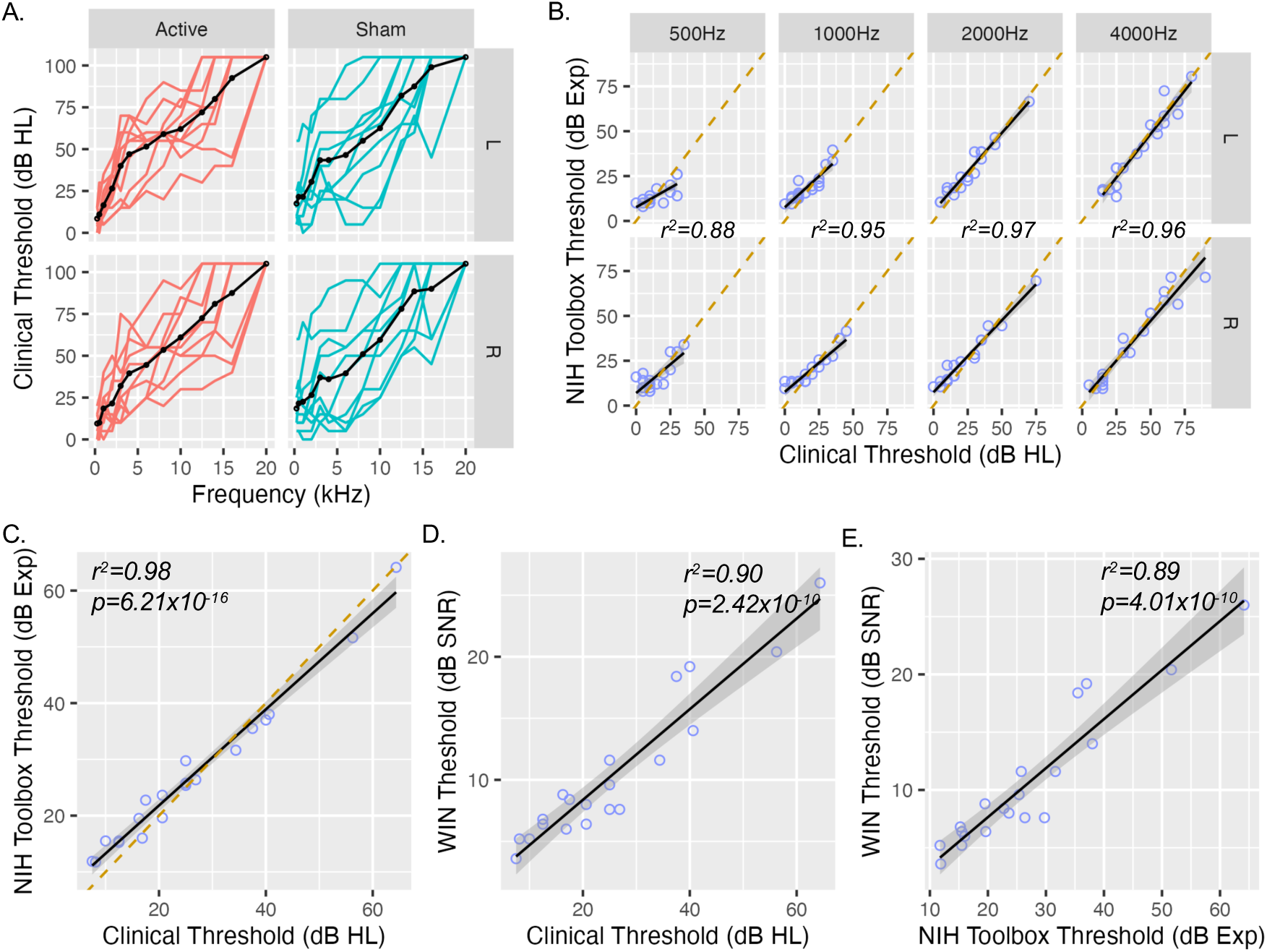
Comparing NIH Toolbox Hearing Assessments and Clinical Audiometry. **A.** Pure tone thresholds tested during clinical audiometry are plotted for standard and extended frequencies for single subjects (red, active group; cyan sham group) and group average (black). **B.** Pure tone thresholds measured during NIH Toolbox Experimental (Exp) testing and clinical audiometry are plotted for each subject (purple circles) at baseline before tDCS. Black lines show linear fit, and orange dashed line shows x=y. R squared is given for each frequency averaged over each ear, all p < 0.000005. **C-E.** Scatter plots demonstrate relationships between mean pure-tone threshold measured using NIH Toolbox and during Clinical Audiometry (matched frequencies; plotted as in B), and threshold on the NIH Toolbox Words in Noise task.

After tDCS, no change was detected in mean experimental pure-tone thresholds measured with NIH Toolbox (**Figure 3A**, **Table 2**). There was no time-by-group interaction or main effect of time for mean threshold, or when separating data by ear tested (all p > 0.15, **Supplemental Table 1**).

**Figure 3.**
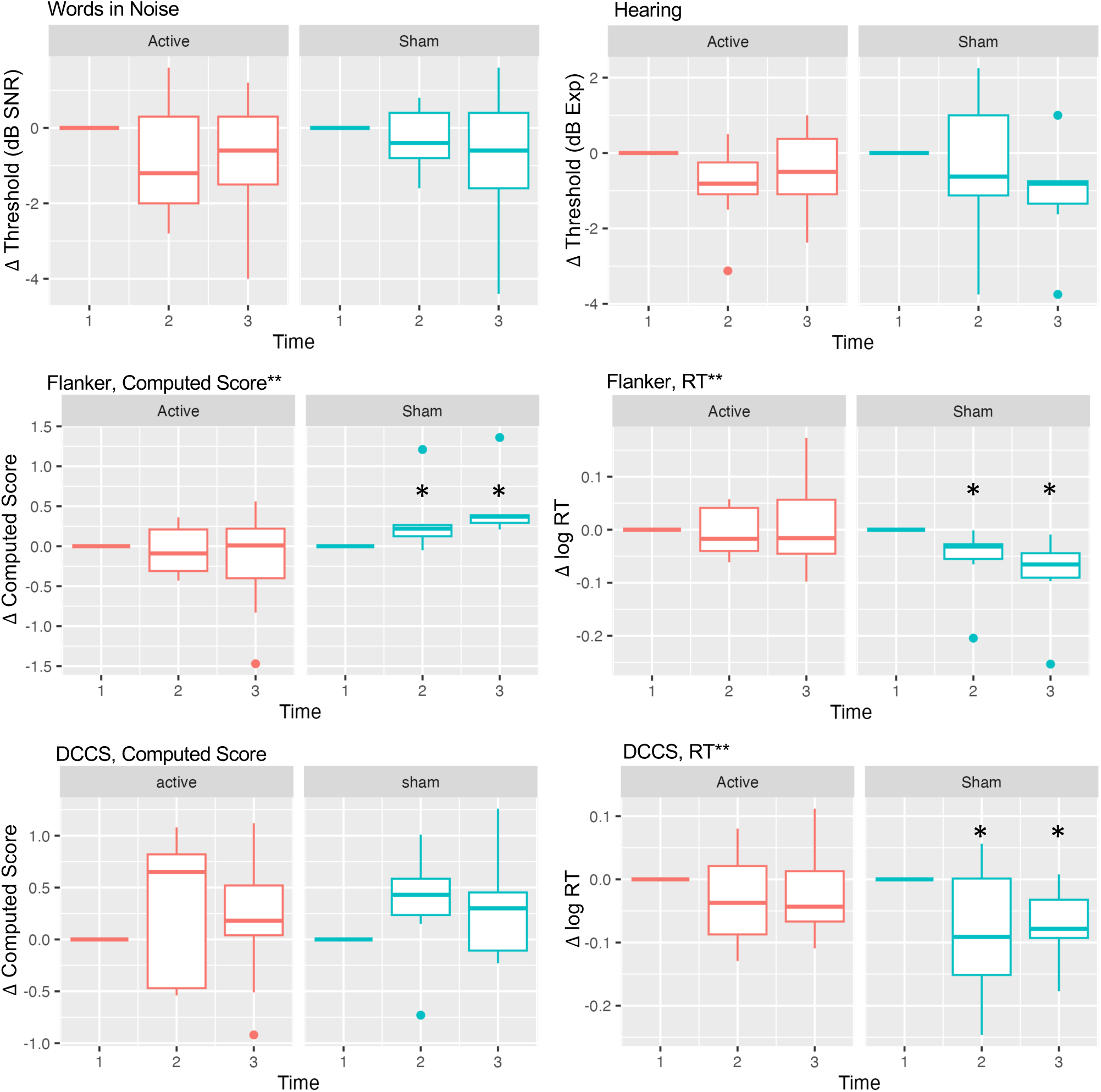
Results of auditory and cognitive tasks before and after focal auditory-cortex tDCS measured with NIH Toolbox. **A.** Boxplots pure-tone thresholds before and after tDCS for each group (Active, Sham) and ear (Left L; Right R), averaged over frequencies tested. **B.** Words in Noise thresholds are plotted for for each group and ear (Left L; Right R), averaged over all SNR levels tested. **C&D.** Change in Computed Score and reaction time (RT) during correct responses on the Flanker Task are plotted for each group. **E&F.** Change in Computed Score and RT are plotted for the Dimension Change Card Sort (DCCS) task. In all plots, Time 1 on x axes represents baseline session before tDCS, Time 2 is immediately after the first tDCS session, and Time 3 is after the fifth tDCS session. Asterisks mark pairwise change from baseline p < 0.05 in cases of significant main effect of time or interaction.

**Table 2.** Auditory task performance after focal auditory cortex tDCS.

| Task | Effect | $\beta$ | $\beta$ SE | df | t | p |
| --- | --- | --- | --- | --- | --- | --- |
| Hearing | Time-by-Group Interaction | -0.27 | 0.63 | 93 | -0.42 | 0.67 |
|  | Time | -0.24 | 0.43 | 93 | -0.56 | 0.58 |
|  | Condition (Ear) | -2.11 | 0.50 | 93 | -4.20 | 0.0001 |
|  | Active, time1 - time2 | 0.84 | 0.86 | 91 | 0.97 | 0.33 |
|  | Active, time1 - time3 | 0.48 | 0.86 | 91 | 0.55 | 0.58 |
|  | Sham, time1 - time2 | 0.26 | 0.90 | 91 | 0.30 | 0.77 |
|  | Sham, time1 - time3 | 1.01 | 0.93 | 91 | 1.08 | 0.28 |
| Word in Noise | Time-by-Group Interaction | 0.04 | 0.38 | 93 | 0.10 | 0.92 |
|  | Time | -0.38 | 0.26 | 93 | -1.47 | 0.15 |
|  | Condition (Ear) | -0.81 | 0.30 | 93 | -2.68 | 0.009 |
|  | Active, time1 - time2 | 0.76 | 0.54 | 92 | 1.41 | 0.16 |
|  | Active, time1 - time3 | 0.76 | 0.54 | 92 | 1.41 | 0.16 |
|  | Sham, time1 - time2 | 0.26 | 0.56 | 92 | 0.46 | 0.65 |
|  | Sham, time1 - time3 | 0.68 | 0.58 | 92 | 1.17 | 0.24 |

Similarly, there was no time-by-group interaction or main effect of time for overall WIN threshold (**Figure 3B**, **Table 2**). However, there was a modest main effect of time for WIN threshold for stimuli presented to the right ear (p=0.043, **Supplemental Table 1**). For right-ear stimuli, WIN threshold decreased for the active group between baseline and the 5^th^ tDCS session (p=0.047). WIN scores also tended to improve after the first active tDCS session and after sham tDCS for right-ear stimuli, though these changes did not reach statistical criterion (all p>0.11, **Supplemental Table 1**). There was no main effect of time for left ear stimuli, and no time-by-group interaction for either ear (**Supplemental Table 1**).

Performance on both auditory tasks was better for stimuli presented to the right ear, compared with stimuli presented to the left (main effect of condition/ear, **Table 2**). However, this overall advantage for right-ear stimuli did not change over time after active or sham tDCS (**Supplemental Table 2**).

In the Flanker task, a time-by-group interaction was noted for Computed Scores, where performance improved after sham but not active tDCS (**Table 3**, **Figure 3**). Computed Score is calculated using both accuracy and RT, and accuracy averaged across all three sessions was near/at ceiling (95% accuracy for one sham volunteer and 100% for all others). Therefore, reaction time (RT) was also analyzed separately. Indeed, a time-by-group interaction was also present for RT, where RTs decreased after the 1^st^ and 5^th^ sham tDCS session, but did not change after active tDCS (**Figure 3C**, **Table 2**). Time-by-group interactions on RT were present for both Congruent trials and Incongruent trials when analyzed separately; RTs were faster for each condition after the final sham tDCS session, and after the first sham tDCS session on Congruent trials (all p<0.01, **Supplemental Table 1**). Again, RTs did not change after active tDCS for Congruent or Incongruent trials (all p>0.5).

**Table 3.**
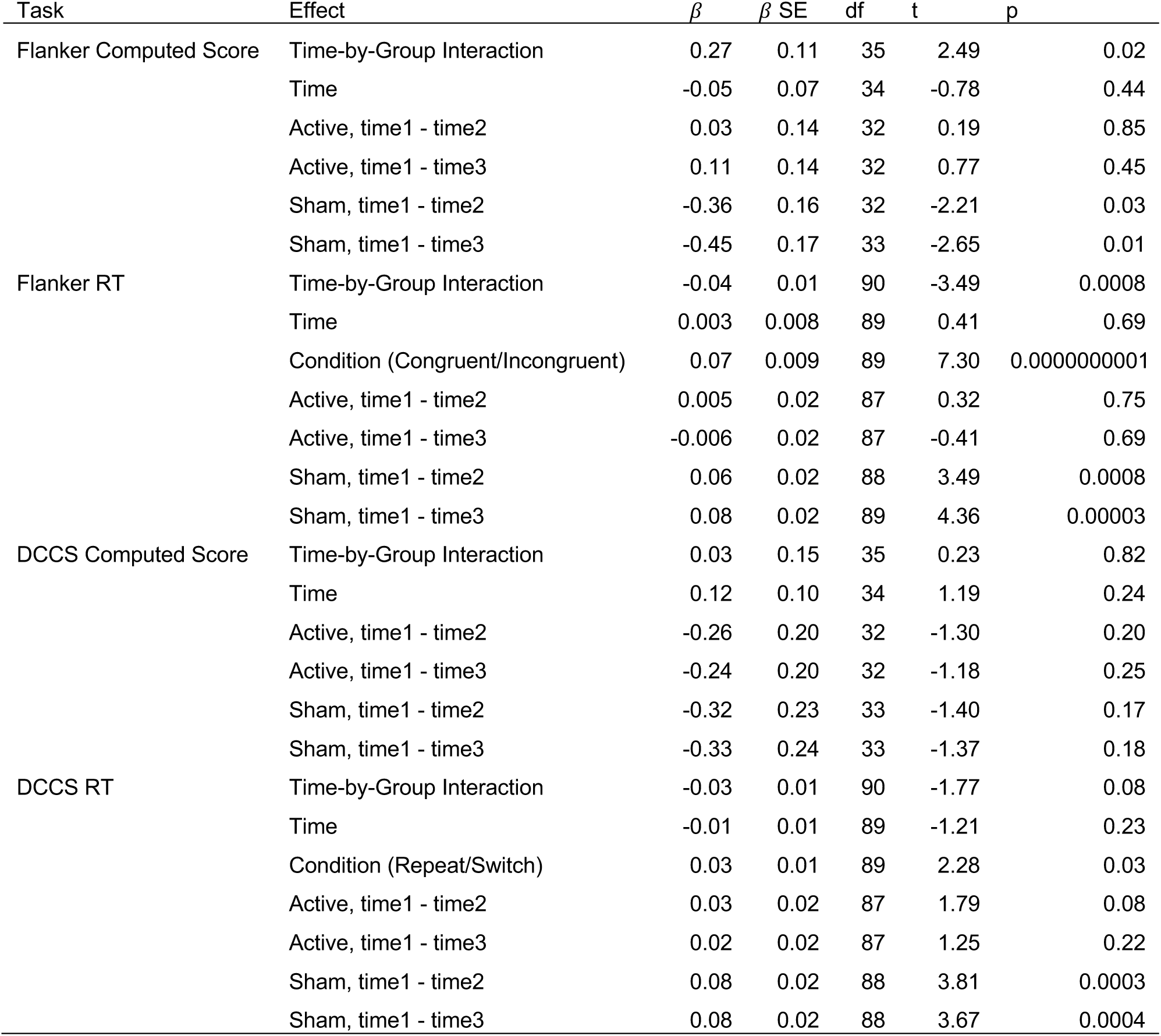
Cognitive task performance after focal auditory-cortex tDCS.

In the DCCS task, no time-by-group interaction or main effect of time was present for Computed Scores (Table 3, Figure 3). Accuracy for the DCCS Task was also high; mean accuracy was 98.3% (SD=0.05%) for the active group and 96.9% (SD=0.08%) for sham with no difference between groups (t(18)=-0.84, p=0.41). However, a time-by-group interaction for DCCS RTs did not reach statistical criterion (p=0.08, **Table 2**, **Figure 3D**), though RTs did decrease after the first and fifth sham tDCS sessions (**Table 2**). When analyzing Repeat and Switch trials separately, the overall pattern of faster RTs after sham tDCS persisted for Repeat trials, but the time-by-group interaction did not reach criterion (p=0.057, **Supplemental Table 1**).

As expected, RTs were lower (faster) on Congruent trials than on Incongruent trials for the Flanker Task, and lower for Repeat than for Switch trials on the DCCS Task (main effect of task condition, **Table 2**). However, this effect (i.e., decreased RTs during Congruent and Repeat trials) did not change over time in either the active or sham groups (**Supplemental Table 2**). Descriptive statistics for all auditory and cognitive measures are listed in **Supplemental Table 3**.

## DISCUSSION

This pilot study demonstrates the feasibility of brief auditory and cognitive testing during clinical trials for tinnitus using a simple tablet-based experimental set up, providing an alternative to clinical audiometry and neuropsychological testing to decrease volunteer burden. Importantly, scores for both clinically and experimentally administered hearing tests were highly correlated before tDCS,^26,27^ suggesting that clinical audiometry may not be needed when measuring potential adverse effects on hearing in these trials. This study also provides evidence that focal auditory-cortex tDCS may influence some aspects of auditory perception and cognition. Repeated stimulation of auditory cortex with focal tDCS did not impair performance on basic auditory tasks in the current study. However, active tDCS seemed to interfere with learning “practice effects” during cognitive tasks, and may have modestly improved hearing in noise. Taken together, these results demonstrate the feasibility and importance of measuring both expected and unexpected effects of brain stimulation on brain function via quick, tablet-based perceptual and cognitive testing.

### Effects of Transcranial Stimulation on Hearing

There is a preponderance of evidence that tDCS can potentiate motor response to TMS, suggesting that tDCS can enhance (or suppress) top-down signaling from motor cortex to effector muscles.^28,29^ From this, one might hypothesize that auditory-cortex tDCS could also modulate bottom-up signaling from hair cells in the inner ear to auditory cortex. Measuring responses to pure-tone stimuli is a mainstay of clinical audiometry and auditory neuroscience, and a sensible way to assess the impact of brain stimulation on bottom-up auditory processing. In the present study, pure-tone thresholds did not change after stimulation, suggesting that the present tDCS protocol does not have long-term beneficial or adverse effects on pure-tone thresholds. However, there is evidence that other stimulation protocols can influence bottom-up auditory processing. For example, transcranial alternating current stimulation (tACS) of auditory cortex can improve detection of simple stimuli (e.g., tones, click trains) presented at positive (and presumably excitatory) phases of electrical current.^30,31^ There are also several studies showing that auditory-cortex tDCS can influence tone-evoked EEG responses after stimulation.^32–35^ Phase-dependent facilitation (and suppression) of tone-evoked EEG responses also occurs with auditory-cortex tACS.^30,36^ Functional MRI data collected during the current study also showed increased cerebral blood flow and functional connectivity in Heschl’s gyrus and surrounding early auditory regions after active auditory-cortex tDCS.^10^ In sum, there is clear evidence that transcranial electrical stimulation can change early auditory cortical function, though auditory-cortex tDCS at standard doses may be insufficient to induce long-term changes in basic pure-tone thresholds.

There is somewhat more evidence that transcranial stimulation can influence complex auditory perception.^37,38^ This could be explained by the fact that stimulation tends to be greater in lateral superior temporal cortex where more complex acoustic features are processed (vs. core or primary auditory areas located more medially^39^), including speech^40^ and detection of speech in noise.^14,16^ In the present study, detection of speech in noise improved for right-ear stimuli after five active tDCS sessions. Average threshold improved 1.2 dB SNR for right-ear stimuli, a very modest effect in relation to estimates of clinically meaningful change (e.g., 6-8 SNR^41^) that should be interpreted with caution. However, the effect is consistent with previous studies. For example, tDCS of left auditory cortex can improve detection of speech and similar sounds in noise,^42–44^ as well as basic perceptual processes relevant to speech perception like auditory stream segregation^45^ and voice onset time.^46^ Studies have also reported that tDCS and tACS can influence performance on gap detection tasks, a common marker of central (vs. peripheral) auditory function.^47–50^ Taken together, the available evidence suggests the potential for NIBS methods like tDCS to modulate central auditory processing.^51,52^ Given that central auditory processing can be negatively impacted in several conditions (e.g., stroke, speech in noise difficulties, auditory processing disorder), this could reflect an untapped area of future translational research.

Though not a focus of the present study, performance on both auditory tasks was better for stimuli presented to the right ear. A right ear advantage for speech perception has been noted previously, and is thought to reflect both the prevalence of left-hemisphere dominance for language and midline crossing of the ascending auditory system.^53,54^ The left ear also appears to be more susceptible to hearing loss assessed with pure tone audiometry,^55–57^ which may explain the right-ear advantage for pure tone detection in the current group of tinnitus volunteers, some of whom had hearing loss. This right-ear advantage did not change over time after active tDCS in either task, which is consistent with negative findings in dichotic listening studies of conventional anodal tDCS^58^ or unilateral transcranial random noise stimulation^59^ of auditory cortex. However, the current study was not designed to measure this, and it is likely that other NIBS protocols targeting auditory cortex can modulate this phenomenon.^59,60^

### Effects on Cognitive Task Performance

The intended stimulation target in the present study was superior temporal (auditory) cortex, and no aspects of cognition were anticipated to be affected. Yet, the present study suggests that anodal tDCS of left auditory cortex interferes with motor learning during cognitive tasks, but not cognitive inhibition (flanker task) or cognitive flexibility (rule switching during DCCS task). After sham tDCS, reaction times were faster after repeat administration of the Flanker and DCCS Tasks, an expected effect reflecting well-known “practice effects” for many cognitive and perceptual tasks. However, reaction times for the active group did not improve over repeated testing, which was unexpected. In previous studies, anodal auditory-cortex tDCS has interfered with learning during pitch/frequency discrimination,^61,62^ yet it is unclear why auditory-cortex tDCS would interfere with this basic form of visuomotor learning during cognitive tasks in the current study.

One possible interpretation of these results is that anodal (positive) current in the ventral motor strip and/or cathodal (negative) current in premotor and/or parietal regions interfered with practice effects on reaction time during cognitive tasks (**Figure 1A**). Indeed, anodal stimulation of dorsal motor cortex near hand/digit representations is strongly associated with improved motor learning.^63,64^ Perhaps anodal stimulation of ventral motor cortex or cathodal stimulation of premotor cortex could disrupt these effects, though note that the amplitude of stimulation per cathode in the present study (0.5 mA) is less than what is typically used in studies showing effects on cognition and perception. Reversing current polarity in the present tDCS protocol (i.e., center cathode, surround anodes) to measure potential improvements in reaction time active stimulation over and above sham stimulation would be informative, and of course independent validation of the present results in a larger sample is needed.

Of note, the current results support the validity of the Flanker and DCCS NIH Toolbox tasks in measuring cognitive inhibition and flexibility.^13^ Reaction times were significantly higher during Incongruent and Switch trials on these tasks, respectively. Though not a focus of the current study, prefrontal stimulation is more likely to influence these aspects of cognitive task performance.^65,66^

### Conclusions

This study provides proof-of-concept evidence for the utility of brief, tablet-based audiometric and cognitive assessments during brain stimulation trials. These ancillary analyses demonstrated the safety of multiple sessions of 4×1 Ag/AgCl tDCS targeting auditory cortex in people with chronic tinnitus and hearing loss. Pure tone and speech reception thresholds did not worsen during this study, and detection of speech in noise may have modestly improved, though this latter effect requires independent validation in a larger sample. Importantly, these experimental thresholds correlated strongly with clinical audiometry, supporting the external and clinical validity of tablet-based experimental audiometry.^26,27^ The present analyses also demonstrate that tablet-based tasks can be used to find unexpected effects of transcranial brain stimulation on perception and cognition. In this study, stimulation targeting auditory cortex interfered with improved RTs over repeat administration of cognitive tasks, perhaps reflecting unintended effects of stimulation “bleeding” into ventral motor, premotor/prefrontal, and/or parietal cortex. Despite these strengths, there are a number of limitations of the present study that should be considered, including the small sample size and lack of control group without chronic tinnitus or hearing loss. This latter point is particularly important given that both tinnitus and hearing loss can affect auditory cortex function,^67,68^ and people without tinnitus or hearing loss might respond differently to this auditory-cortex tDCS protocol. The NIH Toolbox itself necessarily has limitations as well, including low difficulty (ceiling performance accuracy) on Flanker and DCCS tasks, limited range of pure-tone frequencies tested, and inability to track reaction times during auditory assessments. Nevertheless, these limitations are present in many clinical studies, and does not preclude the potential usefulness of the present dataset, the NIH toolbox, or similar approaches. In sum, adding brief, targeted task batteries to future studies and trials could be a low-cost, low-burden way to test for expected and unexpected effects of NIBS and to improve the impact NIBS research.

## Data Availability

All data produced in the present study are available upon reasonable request to the author

## ACKNOWLEDGEMENTS

This work was supported by the National Institutes of Health under award number DC015880 to Dr. Leaver. This content is solely the responsibility of the authors and does not necessarily reflect the official views of the National Institutes of Health.

## CONFLICT OF INTEREST

The author declares no conflicts of interest.

## Notes

Financial Support: National Institutes of Health DC015880

Conflict of Interest Statement: The author reports no conflicts of interest.

### Competing Interest Statement

All authors have completed the ICMJE uniform disclosure form at www.icmje.org/coi_disclosure.pdf and declare: all authors had financial support from National Institutes of Health for the submitted work; no financial relationships with any organizations that might have an interest in the submitted work in the previous three years; no other relationships or activities that could appear to have influenced the submitted work.

### Clinical Trial

NCT05120037

### Author Declarations

Institutional Review Board of Northwestern University gave ethical approval for this work.

