## Supplemental Table for "Perceptual and cognitive effects of focal tDCS of auditory cortex in tinnitus"

Running Title: Cognitive-perceptual effects of focal tDCS in tinnitus

Amber M. Leaver PhD[1]*

[1] Department of Radiology, Northwestern University, Chicago, IL, USA

**Supplemental Table 1, Changes in auditory and cognitive task conditions after tDCS**

| Task, Condition | Effect | beta | beta SE | df | t.ratio | p.value |
|---|---|---|---|---|---|---|
| Hearing, Right Ear | Time-by-Group Interaction | -0.40 | 0.38 | 36 | -1.05 | 0.30 |
| | Time | -0.38 | 0.25 | 36 | -1.47 | 0.15 |
| | Active, time1 - time2 | 0.92 | 0.49 | 34 | 1.89 | 0.07 |
| | Active, time1 - time3 | 0.75 | 0.49 | 34 | 1.53 | 0.13 |
| | Sham, time1 - time2 | -0.02 | 0.51 | 34 | -0.04 | 0.97 |
| | Sham, time1 - time3 | 1.54 | 0.53 | 34 | 2.91 | 0.01 |
| Hearing, Left Ear | Time-by-Group Interaction | -0.17 | 0.51 | 36 | -0.33 | 0.74 |
| | Time | -0.10 | 0.34 | 36 | -0.29 | 0.77 |
| | Active, time1 - time2 | 0.75 | 0.69 | 34 | 1.08 | 0.29 |
| | Active, time1 - time3 | 0.20 | 0.69 | 34 | 0.29 | 0.77 |
| | Sham, time1 - time2 | 0.59 | 0.72 | 34 | 0.82 | 0.42 |
| | Sham, time1 - time3 | 0.54 | 0.75 | 34 | 0.71 | 0.48 |
| Words in Noise, Right Ear | Time-by-Group Interaction | 0.08 | 0.42 | 36 | 0.20 | 0.84 |
| | Time* | -0.60 | 0.29 | 36 | -2.10 | 0.043 |
| | Active, time1 - time2 | 0.56 | 0.58 | 34 | 0.96 | 0.34 |
| | Active, time1 - time3* | 1.20 | 0.58 | 34 | 2.07 | 0.047 |
| | Sham, time1 - time2 | 0.94 | 0.60 | 34 | 1.56 | 0.13 |
| | Sham, time1 - time3 | 1.03 | 0.63 | 34 | 1.64 | 0.11 |
| Words in Noise, Left Ear | Time-by-Group Interaction | 0.01 | 0.54 | 36 | 0.02 | 0.98 |
| | Time | -0.16 | 0.36 | 36 | -0.44 | 0.66 |
| | Active, time1 - time2 | 0.96 | 0.72 | 34 | 1.33 | 0.19 |
| | Active, time1 - time3 | 0.32 | 0.72 | 34 | 0.44 | 0.66 |
| | Sham, time1 - time2 | -0.46 | 0.75 | 34 | -0.61 | 0.55 |
| | Sham, time1 - time3 | 0.29 | 0.78 | 34 | 0.38 | 0.71 |
| Flanker, Congruent | Time-by-Group Interaction* | -0.05 | 0.02 | 35.37 | -2.79 | 0.008 |
| | Time | 0.00 | 0.01 | 34.45 | 0.21 | 0.83 |
| | Active, time1 - time2 | 0.01 | 0.02 | 32.01 | 0.22 | 0.83 |
| | Active, time1 - time3 | 0.00 | 0.02 | 32.01 | -0.22 | 0.83 |
| | Sham, time1 - time2* | 0.08 | 0.03 | 33.04 | 3.17 | 0.003 |
| | Sham, time1 - time3* | 0.10 | 0.03 | 33.58 | 3.64 | 0.0009 |
| Flanker, Incongruent | Time-by-Group Interaction* | -0.04 | 0.01 | 34.54 | -2.37 | 0.02 |
| | Time | 0.00 | 0.01 | 34.17 | 0.41 | 0.69 |
| | Active, time1 - time2 | 0.00 | 0.02 | 32.00 | 0.25 | 0.81 |
| | Active, time1 - time3 | -0.01 | 0.02 | 32.00 | -0.40 | 0.69 |
| | Sham, time1 - time2 | 0.04 | 0.02 | 32.44 | 1.87 | 0.07 |
| | Sham, time1 - time3* | 0.06 | 0.02 | 32.67 | 2.73 | 0.01 |
| DCCS logRT, Repeat | Time-by-Group Interaction | -0.03 | 0.02 | 34.44 | -1.97 | 0.057 |
| | Time | -0.01 | 0.01 | 34.09 | -1.25 | 0.22 |
| | Active, time1 - time2 | 0.03 | 0.02 | 32.00 | 1.60 | 0.12 |

| | | | | | | |
|---|---|---|---|---|---|---|
| | Active, time1 - time3 | 0.03 | 0.02 | 32.00 | 1.29 | 0.21 |
| | Sham, time1 - time2 | 0.08 | 0.02 | 32.36 | 3.33 | 0.002 |
| | Sham, time1 - time3 | 0.10 | 0.03 | 32.56 | 3.89 | 0.0005 |
| DCCS logRT, Switch | Time-by-Group Interaction | -0.02 | 0.02 | 35 | -0.67 | 0.51 |
| | Time | -0.01 | 0.02 | 34 | -0.57 | 0.57 |
| | Active, time1 - time2 | 0.03 | 0.03 | 32 | 1.00 | 0.32 |
| | Active, time1 - time3 | 0.02 | 0.03 | 32 | 0.58 | 0.57 |
| | Sham, time1 - time2 | 0.08 | 0.04 | 33 | 2.11 | 0.04 |
| | Sham, time1 - time3 | 0.06 | 0.04 | 33 | 1.55 | 0.13 |

*Pairwise tests discussed when main effect of time or interaction was $p < 0.05$, but all are listed here for completeness

**Supplemental Table 2, Difference in performance between task conditions does not change over time**

| Task | Effect | beta | beta SE | df | t.ratio | p.value |
|---|---|---|---|---|---|---|
| Hearing, Right vs. Left Ear | Time-by-Group Interaction | -0.02 | 0.03 | 36 | -0.72 | 0.48 |
| | Time | -0.01 | 0.02 | 36 | -0.55 | 0.59 |
| Words in Noise, Right vs. Left Ear | Time-by-Group Interaction | -0.002 | 0.05 | 35 | -0.04 | 0.97 |
| | Time | -0.002 | 0.04 | 35 | -0.06 | 0.95 |
| Flanker, Congruent vs. Incongruent | Time-by-Group Interaction | -0.50 | 0.37 | 52 | -1.34 | 0.19 |
| | Time | -0.10 | 0.25 | 52 | -0.41 | 0.68 |
| DCCS, Repeat vs. Switch | Time-by-Group Interaction | -1.85 | 1.30 | 52 | -1.42 | 0.16 |
| | Time | -0.14 | 0.87 | 52 | -0.16 | 0.87 |

**Supplemental Table 3, Task performance across tSDCS sessions**

| | | | Mean | | | SD | | |
|---|---|---|---|---|---|---|---|---|
| Task Measure | Condition | Group | Time 1 | Time 2 | Time 3 | Time 1 | Time 2 | Time 3 |
| Hearing Threshold, dB Exp | Right | Active | 24.25 | 23.33 | 23.50 | 9.37 | 9.40 | 9.22 |
| | | Sham | 23.24 | 27.55 | 27.63 | 13.82 | 18.57 | 19.15 |
| | Left | Active | 26.63 | 25.88 | 26.43 | 8.45 | 8.37 | 8.32 |
| | | Sham | 25.21 | 28.40 | 29.57 | 11.93 | 16.44 | 16.27 |
| Word in Noise, dB SNR | Right | Active | 9.12 | 8.56 | 7.92 | 3.91 | 4.74 | 4.71 |
| | | Sham | 10.00 | 10.72 | 11.07 | 5.20 | 7.58 | 8.02 |
| | Left | Active | 10.24 | 9.28 | 9.92 | 4.89 | 4.26 | 3.40 |
| | | Sham | 9.56 | 11.60 | 11.51 | 6.92 | 7.90 | 7.82 |
| Flanker Computed Score | N/A | Active | 8.38 | 8.36 | 8.28 | 0.32 | 0.48 | 0.69 |
| | | Sham | 7.93 | 8.38 | 8.45 | 1.09 | 1.00 | 1.01 |
| Flanker RT, sec | Congruent | Active | 0.65 | 0.64 | 0.67 | 0.10 | 0.12 | 0.16 |
| | | Sham | 0.72 | 0.60 | 0.57 | 0.20 | 0.14 | 0.15 |
| | Incongruent | Active | 0.73 | 0.73 | 0.76 | 0.10 | 0.13 | 0.18 |
| | | Sham | 0.88 | 0.77 | 0.72 | 0.45 | 0.36 | 0.32 |
| DCCS Computed Score | N/A | Active | 8.19 | 8.45 | 8.42 | 0.58 | 0.77 | 0.82 |
| | | Sham | 8.05 | 8.15 | 8.11 | 1.55 | 1.40 | 1.11 |
| DCCS RT, sec | Repeat | Active | 0.72 | 0.67 | 0.68 | 0.16 | 0.19 | 0.19 |
| | | Sham | 0.80 | 0.73 | 0.67 | 0.37 | 0.37 | 0.21 |
| | Switch | Active | 0.72 | 0.68 | 0.71 | 0.14 | 0.19 | 0.22 |
| | | Sham | 0.86 | 0.81 | 0.80 | 0.56 | 0.47 | 0.37 |
